# Exhaled breath is a significant source of SARS-CoV-2 emission

**DOI:** 10.1101/2020.05.31.20115154

**Authors:** Jianxin Ma, Xiao Qi, Haoxuan Chen, Xinyue Li, Zheng Zhang, Haibin Wang, Lingli Sun, Lu Zhang, Jiazhen Guo, Lidia Morawska, Sergey A. Grinshpun, Pratim Biswas, Richard C. Flagan, Maosheng Yao

## Abstract

Despite notable efforts in airborne SARS-CoV-2 detection, no clear evidence has emerged to show how SARS-CoV-2 is emitted into the environments. Here, 35 COVID-19 subjects were recruited; exhaled breath condensate (EBC), air samples and surface swabs were collected and analyzed for SARS-CoV-2 using reverse transcription-polymerase chain reaction (RT-PCR). EBC samples had the highest positive rate (16.7%, n = 30), followed by surface swabs(5.4%, n = 242), and air samples (3.8%, n = 26). COVID-19 patients were shown to exhale SARSCoV-2 into the air at an estimated rate of 10^3^-10^5^ RNA copies/min; while toilet and floor surfaces represented two important SARS-CoV-2 reservoirs. Our results imply that airborne transmission of SARS-CoV-2 plays a major role in COVID-19 spread, especially during the early stages of the disease.

**One Sentence Summary:** COVID-19 patient exhales millions of SARS-CoV-2 particles per hour

---

The COVID-19 pandemic has left a major mark on human history. Global efforts to intervene the spread are accelerating, however the knowledge on the major routes of COVID-19 transmission is required. Analysis of environmental samples provides clues *(1–4)*. Notably, SARS-CoV-2 has been detected in air *(2–4)*, on ventilation fans *(1)* and hospital floors *(1,4)*. Surface swabs from keyboards, cell phones, and patients’ hands have also tested positive *(1)*. Other studies have shown that aerosolized SARS-CoV-2 not only survives on various surfaces for sustained periods of time *(5)*, but also remains viable in the air for up to 3 hours *(6)*. Despite these rapid developments, the key COVID-19 transmission routes still remain debated *(7)*, and evidence is extremely sparse on how SARS-CoV-2 is emitted into the air. We investigated the hypothesized transmission routes of SARS-CoV-2 by examining 35 enrolled COVID-19 patients (both imported and local) (Table S1) in Beijing. A total of 298 samples were collected, including 30 exhaled breath condensate (EBC), 26 air and 242 surface swab samples. Our data reveal direct evidence of airborne transmission of SARS-CoV-2 via breathing.

SARS-CoV-2 RNA was detected in exhaled breath from five COVID-19 patients aged under 50 (Fig. 1, Table S1). The times from symptom onset to the EBC collection were all less than 14 days (Fig. 1A). Surface swabs from the cell phone and hands of one patient(ITA-YL1) tested negative for the virus, but the SARS-CoV-2 was present on the toilet pit surface in that patient’s hotel room (Fig. 1C). SARS-CoV-2 was found on another patient’s (UK-YY1) cell phone. Although EBC samples from two patients (ITA-YW2 and UK-YJ1) were shown to contain SARS-CoV-2, surface swabs from their cell phones, hands, and toilet surfaces were negative for the virus. In the ward of patient BJ-YZ3, the virus was present on the surface of an air ventilation duct entrance that was located below the patient’s bed (Video S2). This patient had contracted the COVID-19 from a family member (US-YW1) who had returned from overseas. Cycle threshold (Ct) values were obtained for each positive sample (Fig. 1D and Table S2). The Ct values for EBC samples varied greatly among the patients, with lower values detected for earlier disease stages (Fig. 1D). The overall SARS-CoV-2 positive rate for EBC samples was 16.7% (n = 30). The exhaled SARS-CoV-2 could be partially responsible for the contamination on the surfaces that was observed.

**Fig. 1.**
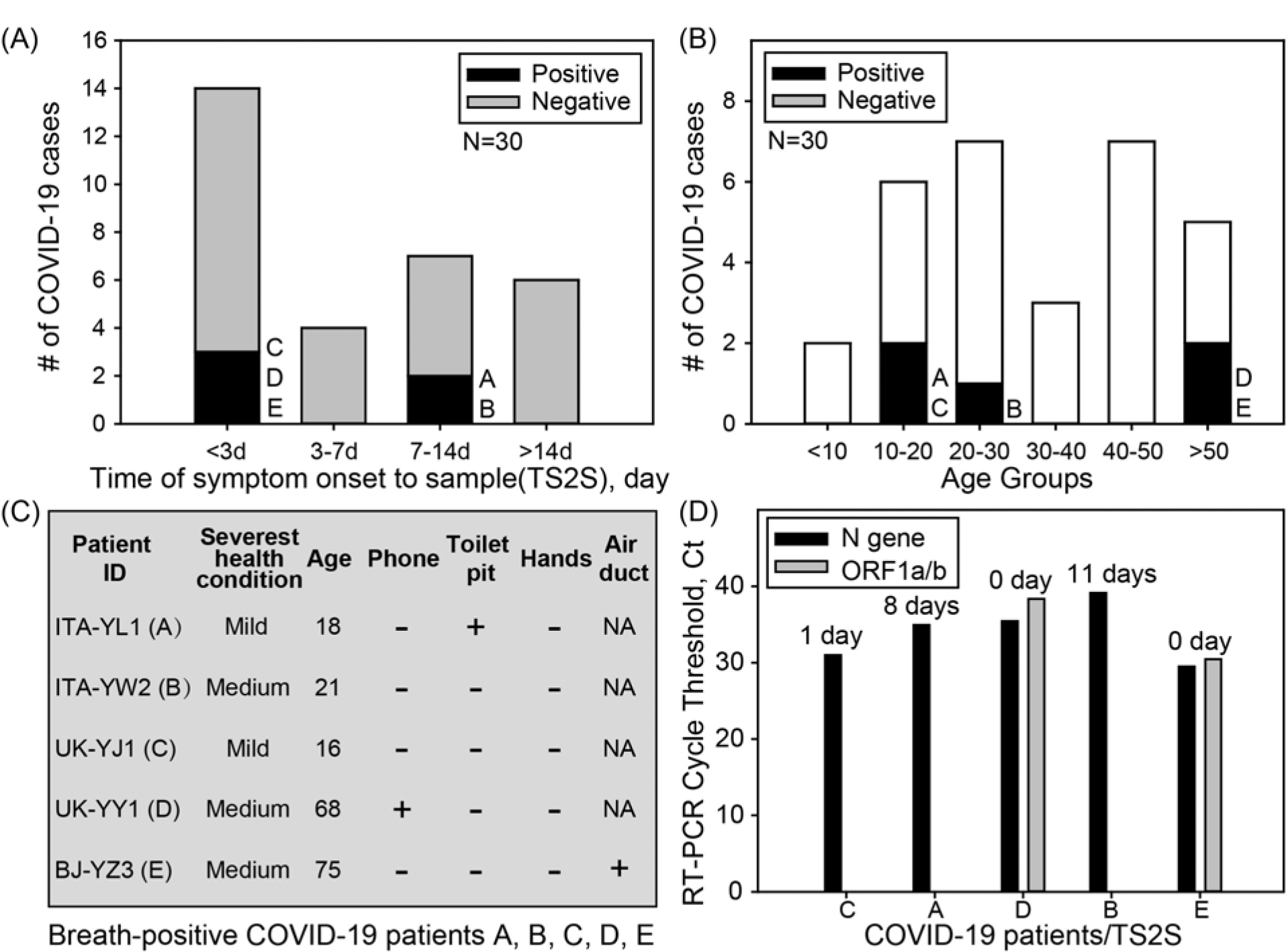
Detection of SARS-CoV-2 from EBC samples collected from 27 COVID-19 patients. A) Number of EBC samples vs Time of Symptom to Sample(TS2S); B) Number of EBC samples by different age groups; C) Details of patients with positive EBC samples; D) RT-PCR cycle threshold (Ct) values of EBC samples with different TS2S values for patients (A, B, C, D, E).

From 26 air samples collected including those using a robot (Video S1), one sample (20YJ3368) from an unventilated quarantine hotel toilet room was positive (Fig. 2A, Table S3). Three people from one family (TH-YZ1, TH-YY1 and TH-YY11) returning from an overseas trip shared the same hotel room. The two family members (TH-YZ1 and TH-YY1) were infected, but the two-year-old child (TH-YY11) was free from the virus. Surface swab samples from a pillow case (20YJ3372) and hands (20YJ3378) of patient TH-YZ1 were shown to contain SARS-CoV-2, but no virus was detected in this patient’s EBC sample (Fig. 2B). Additionally, SARS-CoV-2 was detected on an air ventilation duct entrance surface as described above (the duct acted like an air sampler) (Fig. 1C, Fig. 2C). Overall, 3.8% (n = 26) of air samples were positive for SARS-CoV-2. These air sample data, despite the low positive rate, still show that the air spaces of the hospitals housing the COVID-19 patients were contaminated with SARS-CoV-2.

**Fig. 2.**
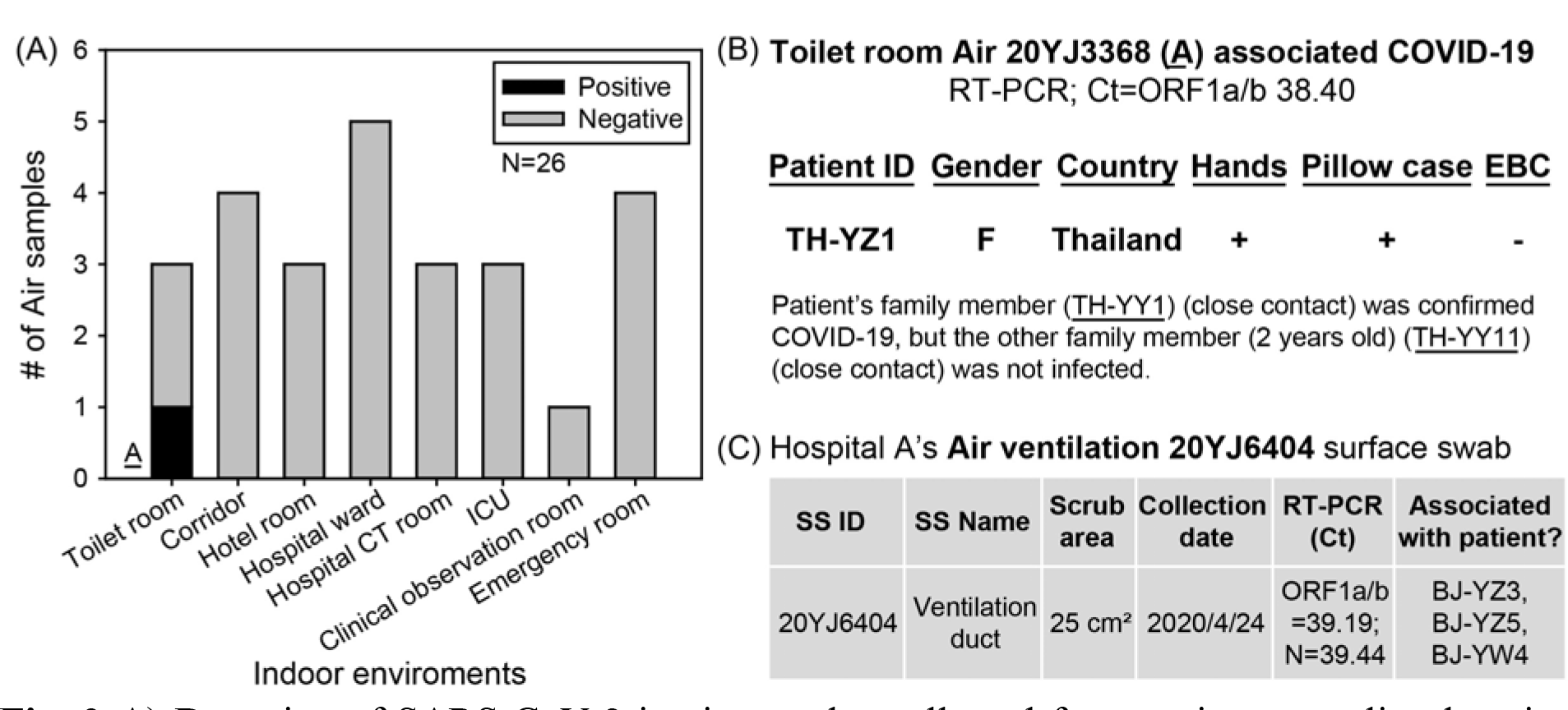
A) Detection of SARS-CoV-2 in air samples collected from various sampling locations; B) patient information associated with the positive toilet sample; C) details of a SARS-CoV-2 positive sample from an air ventilation duct entrance in Hospital A.

Out of 242 surface swab samples, 13 were positive for SARS-CoV-2 (Fig. 3 and Table S4). Among the five categories of surfaces, the Toilet pit had the highest SARS-CoV-2 positive rate(16.7%, n = 12), followed by the Hospital floor (12.5%, n = 16), the Other surfaces (7.4%, n = 27), the Patient touching surfaces (4.0%, n = 149), and the Medical touching surfaces (2.6%, n = 38). Two positive toilet pit samples (20YJ2545, 20YJ2661) were associated with COVID-19 patients (BJ-YC1, ITA-YL1) (Table S1, Fig. 3B). For toilet pit swab 20YJ2661, the EBC (20YJ2771) of its associated patient (ITA-YL1) also tested positive. Another two positive swab samples were from the Hospital floor (Diagnosis room floor- 20YJ2972) and the Clinical observation room (seat pedal- 20YJ3029) (Table S4). For the Patient touching surfaces group, we detected six positive samples from hands (TH-YZ1), a pillow case (TH-YZ1), mobile phones (UK-YY1, USYW1), and computer keyboards (US-YW1, BJ-YW4) (Fig. 3B). In addition, we detected one positive sample(20YJ2757) from the patient transport cart in the computed tomography (CT) room (the Medical touching surfaces), one positive sample (20YJ3010) from the handrail of the clinic corridor, and another (20YJ6404) from the air ventilation duct entrance. Surprisingly, only 2 out of 22 surface swabs from the mobile phones of COVID-19 patients tested positive (Fig. 3B), with a positive rate of approximately half that of the EBC samples (16.7%, n = 30). None of the 26 surface swabs collected from handles of various objects appeared positive for the virus (Table S4). The overall SARS-CoV-2 positive rate for the surface swabs was 5.4% (n = 242). These observations do not support the widely-held belief that direct transmission by contact with surfaces plays a major role in COVID-19 spread.

**Fig. 3.**
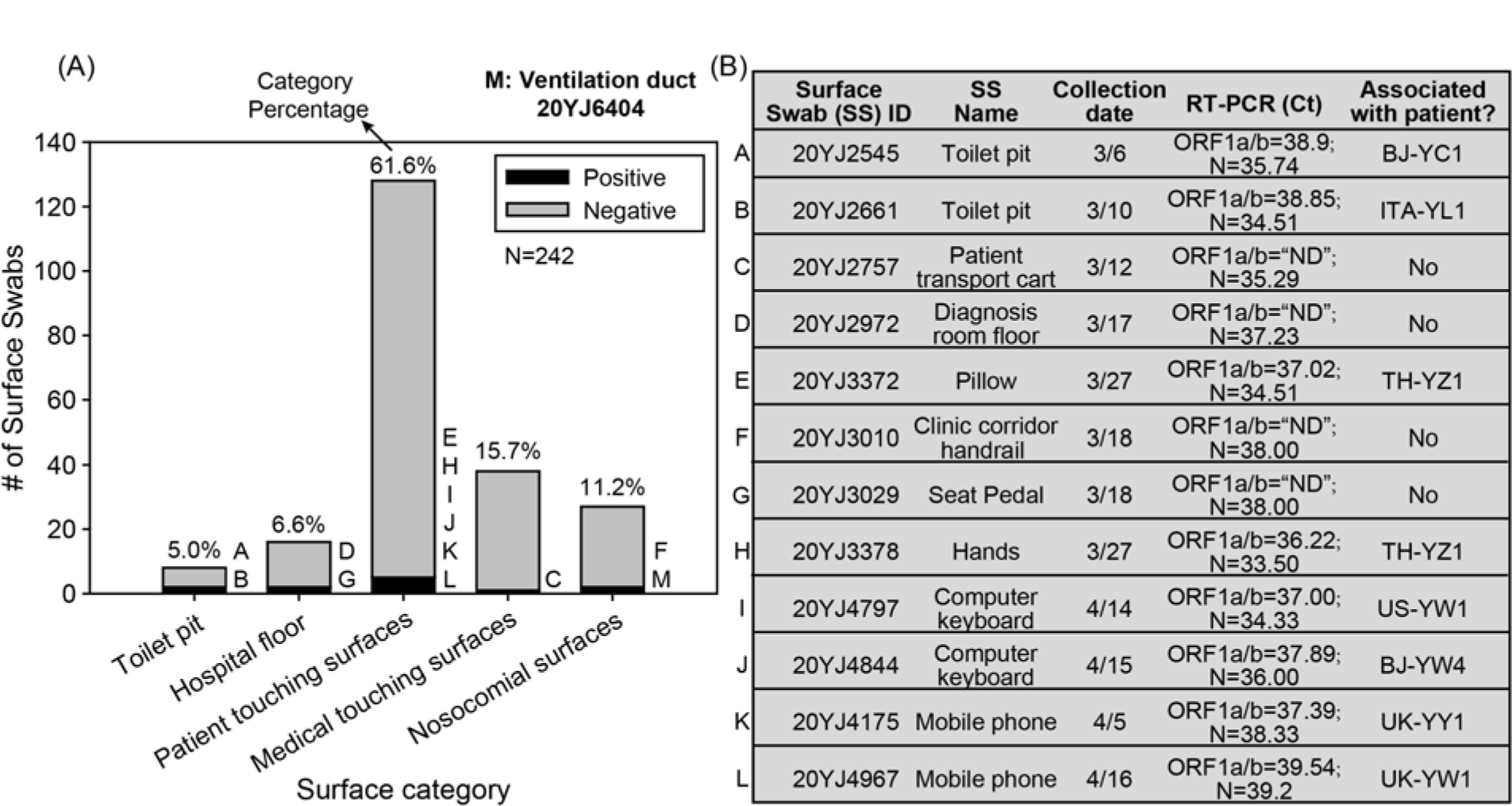
Detection of SARS-CoV-2 in surface swabs from five sampling locations: A) distribution statistics for both positive and negative samples in five surface groups; B) associated details for the detected positive surface samples.

For the first time, we here report that the SARS-CoV-2 is released directly into the air via breathing by COVID-19 patients. The detection limit for the RT-PCR for SARS-CoV-2 detection was reported to be approximately 500 RNA copies per reaction *(8)*. Assuming an amplification efficiency of 75%, the breath emission rate of SARS-CoV-2 was estimated to be about10^3^-10^5^ RNA copies/min. The observed Ct values suggest that SARS-CoV-2 levels in exhaled breath could reach 10^5^-10^7^ copies/m^3^ if an average breathing rate of 12 L/min is assumed. The SARS-CoV-2 emission rate is affected by many factors such as disease stage, patient activity, and, possibly, age. We found that the SARS-CoV-2 breath emission rate into the air was the highest, up to 10^5^ viruses per min, during the earlier stages of COVID-19. This finding was consistent with a previous report that the highest SARS-CoV-2 load in throat swabs was observed at the time of symptom onset *(9)*. Another significant discovery from this work is that SARS-CoV-2 emission was not, however, continuous at the same rate, but was rather a sporadic event. For example, two EBC samples (20YJ2640,20YJ2771) collected from the same patient (ITA-YL1), but on different dates, using the same method returned different test results (Table S2).

SARS-CoV-2 has previously been detected in fine particles in hospital air *(4)*. In public environments such as a classroom and a subway, fine bioaerosol particles with a peak of around 1 μm were detected using a fluorescence-based sensor; their concentration levels were substantially higher than those of the coarse ones *(10,11)*. Most SARS-CoV-2 in exhaled breath should fall in the fine aerosol size (<2.5 μm) ranges, which can remain airborne for far longer time than the coarser ones emitted otherwise in a sneeze or cough. The negative SARS-CoV-2 results (Fig. 2) may result from low SARS-CoV-2 emissions, virus inactivation by disinfectants, and rapid dilution or wash-out of SARS-CoV-2 by fresh air (2.5 m^3^/min, Video S2). The spread of COVID-19 by asymptomatic patients has been also documented *(12,13)*. The asymptomatic disease carriers do not, generally, cough or sneeze to generate respiratory droplets; thus, the observed transmission of the disease has been difficult to explain by respiratory droplet transmission, but is rather logical for a fine aerosol route.

The dominant SARS-CoV-2 transmission routes need to be intervened in order to effectively stop the ongoing COVID-19 pandemic. Large respiratory droplets and direct contact transmissions are presently cited as major transmission routes for the COVID-19. In contrast, we show that the surfaces of mobile phones (n = 22) and various handles (n = 35) frequently used by COVID-19 patients presented very low probabilities of SARS-CoV-2 presence (9.0% and 0%, respectively). Airborne transmission of SARS-CoV-2 has already played an important role in documented real-life COVID-19 spread in semi-enclosed environments *(7,14)*; for example, cluster infection incidents in a choir in Washington State, USA *(15)*, and a restaurant in Guangzhou, China *(16)*. Evidences from our work show that exhaled breath emission may well be the most significant SARS-CoV-2 shedding mechanism, which could have contributed largely to the observed cluster infections and the ongoing pandemic. Accordingly, measures such as enhanced ventilation and the use of face masks are essential to minimize the risk of infection by airborne SARS-CoV-2.

## Funding

This research was supported by a National Natural Science Foundation of China (NSFC)grant (22040101) (PI : M Yao) dedicated to the COVID-19 pandemic. This work was also partially supported by the NSFC Distinguished Young Scholars Fund Awarded to M. Yao (21725701).

## Author contributions

M.Y., and J.M. contributed to the study design. J.M., M.Y., X.Q., Z.Z., H.W., L.S., and J.G., contributed to sample collection and experiments. J.M., X.Q., M.Y. contributed to patients’ recruitment, and clinical management. M.Y., J.M, X.L., H.C., L.Z., L.M., S.A.G., P.B., and R.C.F. contributed to data analysis, data interpretation, figure preparation and literature search. M.Y. wrote the manuscript draft, and all authors revised the manuscript. All authors reviewed and approved the final version of the report.;

## Competing interests

The authors declare no competing interests.

## Data and materials availability

All data are available in the main text or the supplementary materials.

## Materials and Methods *Statistics of COVID-19 patients*

We recruited a total of 39 subjects, including 35 patients with COVID-19 and four without the disease from Hospital A and Hospital B (Table S1). EBC samples were collected from 20 imported COVID-19 patients from Canada, France, Iran, Italy, Japan, Spain, Thailand, United Kingdom, United States and 7 cases from Beijing (Fig. S1A, Table S2). Sixty-one percent of the COVID-19 patients were aged under 40 (Fig. S1B), and 40% of all COVID-19 patients had mild symptoms (Fig. S1C). Fig. S1D and Fig. S2 show the intensive care unit (ICU) and general ward floor settings of Hospital A, respectively. Medical records from the patients were also obtained at the time of sample collection (Table S1). The ethics involving human subjects including the noninvasive collection of exhaled breath condensate samples was waived due to the urgency of the infectious disease outbreak, and approved by the Ethics Committee of the Center for Disease Control and Prevention of Chaoyang District of Beijing.

### Exhaled breath condensate sample collection

EBC samples were collected from 27 COVID-19 patients using a BioScreen device (Beijing dBlueTech Inc., Beijing, China). The device, which utilizes a hydrophobic film equipped with a cooling module, was previously shown to efficiently collect influenza viruses from exhaled breath *(1)*. Using the same protocol, COVID-19 patients were instructed to exhale for 5 min towards the cooled hydrophobic film through a long straw (Fig. S3A); the exhaled breath from the patients was immediately condensed into tiny droplets (Fig. S3B). The EBC droplets were further collected by rolling a drop of 10 μL of deionized (DI) water across the film surface, allowing the droplets to be scavenged into the DI water droplet. EBC samples of 300–500 μL were collected, immediately pipetted into a Corning tube, and transported to the laboratory for SARS-CoV-2 analysis on the same day. For three COVID-19 patients, EBC samples were collected twice to investigate the differences between samples collected on different dates. A total of 30 EBC samples were collected (Table S2).

### Air sample collection

Air samples were taken from hospital and quarantine hotel environments in Beijing from different sampling locations: the Corridor, the Hotel room, the Hospital CT room, the ICU room, the Toilet room, the Emergence room, the Clinical observation room, and the Hospital ward. The hotel and hospital corridors studied here were provided with fresh air by opening windows or using negative pressure ventilation systems (Fig. S1D). The air samples were collected into 3 mL virus culture liquid (MT0301) (Yocon Biology Inc., Beijing, China) using two impingers (WA-15, WA-400; Beijing dBlueTech, Inc) (Fig. S4). The WA-15 was employed with a sampling flow rate of 15 L/min for enclosed environments such as toilets or ICU rooms; while the WA-400, assisted by a robot, was operated at a 400 L/min flow rate in corridor spaces or ventilated environments (Fig. S4, Video S1). After each sampling, 1.5–2 mL collection liquid remained due to the evaporation. The collected air samples were transported to the laboratory for SARS-CoV-2 analysis on the same day. A total of 26 air samples were obtained (Table S3).

## Surface swab sample collection

The surfaces (10 or 25 cm^2^) of objects in quarantine hotels and hospitals or personal items from tCOVID-19 patients were scrubbed using a wet cotton swab. Upon the collection, surface swab samples were immediately deposited into a 3 mL virus collection liquid (Yocon Biology, Inc), and further delivered to the laboratory for SARS-CoV-2 analysis on the same day. The obtained surface swab samples were divided into five groups according to their sampling locations: the Toilet pit, the Hospital floor, the Patient touching surfaces (those frequently touched by the patients), the Medical touching surfaces (those frequently touched by the medical staff), and Other surfaces (those shared by patients, medical staff or visitors). A total of 242 surface swabs were obtained (Table S4).

### SARS-CoV-2 analysis using reverse transcription polymerase chain reaction (RT-PCR) and *loop-mediated isothermal amplification (LAMP)*

For all collected samples, several steps were carried out for SARS-CoV-2 analysis. First, a 200 μL sample liquid was used to extract SARS-CoV-2 RNA using a MagMAX™ Multi-Sample 96-Well RNA Isolation Kit (Thermo Fisher Scientific, Waltham, MA) according to the manufacturer’s instructions. The final solution volume from the above extraction procedure was 20–30 μL. Then, detection of SARS-CoV-2 targeting both ORF1ab and N genes using a detection kit (Jiangsu Bioperfectus Technologies, Nanjing, China) were performed by RT-PCR (Roche 96 fluorescence qPCR instrument, Roche Molecular Systems, Inc., Pleasanton, CA) under the following cycle conditions: 50°C for 10min, and 97°C for 1min, followed by 45 cycles of 97°C for 5s, and 58°C for 30s. The reaction mixture included 7.5 μL of nucleic acid amplification mix, 5μL of Taq EnzymeMix,4 μL of SARS-CoV-2 reaction mix, 3.5μL of RNA free H_2_O, and 5 μL of sample. DI water was used as a negative control. In accordance with the detection kit’s instructions, samples with Ct values of less than 37 or those detected with a Ct value of 37–40 along with an *“S”* shaped amplification curve were considered positive. As a quality control, SARS-CoV-2 detection was also performed on some of the collected samples using a LAMP chip device (Beijing CapitalBio Technology Co., Ltd., Beijing, China) following the manufacturer’s instructions. For all samplings, one-time-use consumables were used, and DI water served as negative controls.

## Notes

### Competing Interest Statement

The authors have declared no competing interest.

### Author Declarations

The ethics involving human subjects including the non-invasive collection of exhaled breath condensate samples was waived due to the urgency of the infectious disease outbreak, and approved by the Ethics Committee of the Center for Disease Control and Prevention of Chaoyang District of Beijing.

## References and Notes

1. S. W. X. Ong, Y. K. Tan, P. Y. Chia, T. H. Lee, O. T. Ng, M. S. Y. Wong, K. Marimuthu, Air surface environmental, and personal protective equipment contamination by severe acute respiratory syndrome coronavirus 2 (SARS-CoV-2) from a symptomatic patient. JAMA 323, 1610–1612 (2020).

2. Z. D. Guo, Z. Y. Wang, S. F. Zhang, X. Li, L. Li, C. Li, Y. Cui, R. B. Fu, Y. Z. Dong, X. Y. Chi, M. Y. Zhang, K. Liu, C. Cao, B. Liu, K. Zhang, Y. W. Gao, B. Lu, W. Chen, Aerosol and surface distribution of severe acute respiratory syndrome coronavirus 2 in hospital wards, Wuhan, China, 2020. Emerg. Infect. Dis. 10.3201/eid2607.200885 (2020).

3. J. L. Santarpia et al., https://doi.org/10.1101/2020.03.23.20039446 (2020).

4. Y. Liu, Z. Ning, Y. Chen, M. Guo, Y. Liu, N. K. Gali, L. Sun, Y. Duan, J. Cai, D. Westerdahl, X. Liu, K. Xu, K. Ho, H. Kan, Q. Fu, K. Lan, Aerodynamic analysis of SARS-CoV-2 in two Wuhan hospitals. Nature 10.1038/s41586–020–2271–3 (2020).

5. A. Chin, J. Chu, M. Perera, K. Hui, H. L. Yen, M. Chan, M. Peiris, L. Poon, Stability of SARS-CoV-2 in different environmental conditions. Lancet Microbe 1, e10 (2020).

6. N. van Doremalen, T. Bushmaker, D. H. Morris, M. G. Holbrook, A. Gamble, B. N. Williamson, A. Tamin, J.L. Harcourt, N.J. Thornburg, S.I. Gerber, J. O. Lloyd-Smith, E. de Wit, V.J. Munster, Aerosol and surface stability of SARS-CoV-2 as compared with SARS-CoV-1. N. Engl. J. Med. 382, 1564–1567 (2020).

7. K.A. Prather, C.C. Wang, R.T. Schooley, Reducing transmission of SARS-CoV-2. Science 10.1126/science.abc6197 (2020).

8. C. B. F. Vogels et al., https://doi.org/10.1101/2020.03.30.20048108 (2020).

9. X. He, E. H. Y. Lau, P. Wu, X. Deng, J. Wang, X. Hao, Y. C. Lau, J. Y. Wong, Y. Guan, X. Tan, X. Mo, Y. Chen, B. Liao, W. Chen, F. Hu, Q. Zhang, M. Zhong, Y. Wu, L. Zhao, F. Zhang, B. J. Cowling, F. Li, G. M. Leung. Temporal dynamics in viral shedding and transmissibility of COVID-19. Nat. Med. 26, 672–675 (2020).

10. H. Fan, X. Li, J. Deng, G. Da, E. Gehin, M. Yao, Time-dependent size-resolved bacterial and fungal aerosols in Beijing subway. Aerosol Air Qual. Res. 17, 799–809 (2017).

11. C. Xu, C. Wu, M. Yao, Fluorescent bioaerosol particles resulting from human occupancy with and without respirators. Aerosol Air Qual. Res. 17, 198–208 (2017).

12. C. Rothe, M. Schunk, P. Sothmann, G. Bretzel, G. Froeschl, C. Wallrauch, T. Zimmer, V. Thiel, C. Janke, W. Guggemos, M. Seilmaier, C. Drosten, P. Vollmar, K. Zwirglmaier, S. Zange, R. Wölfel, M. Hoelscher, Transmission of 2019-nCoV infection from an asymptomatic contact in Germany. N. Engl. J. Med. 382, 970–971 (2020).

13. X. Pan, D. Chen, Y. Xia, X. Wu, T. Li, X. Ou, L. Zhou, J. Liu, Asymptomatic cases in a family cluster with SARS-CoV-2 infection. Lancet Infect. Dis. 20, 410–411 (2020).

14. National Research Council, “Rapid expert consultation on the possibility of bioaerosol Spread of SARS-CoV-2 for the COVID-19 pandemic (April 1, 2020)” (2020).

15. L. Hamner, P. Dubbel, I. Capron, A. Ross, A. Jordan, J. Lee, J. Lynn, A. Ball, S. Narwal, S. Russell, D. Patrick, H, Leibrand. High SARS-CoV-2 attack rate following exposure at a choir practice—Skagit County, Washington, March 2020. MMWR Morb. Mortal. Wkly. Rep. 69, 606–610 (2020).

16. Y. Li et al., https://doi.org/10.1101/2020.04.16.20067728 (2020).

## References and Notes

1. F. Shen, J. Wang, Z. Xu, Y. Wu, Q. Chen, X Li, J. Xu, L. Li, M. Yao, X. Guo, T. Zhu, Rapid 40 flu diagnosis using silicon nanowire sensor. Nano Lett. 12, 3722-3730(2012).

